# Spatial heterogeneity in *Onchocerca volvulus* IgG4 seroprevalence around a blackfly (*Simulium damnosum* s.l.) breeding site in Ghana and its implications for onchocerciasis serosurveillance

**DOI:** 10.64898/2026.05.05.26352446

**Authors:** Irene Kyomuhangi, Kenneth B. Otabil, Anabel Acheampong, Dennis K. Ofori, Prince-Charles Kudzordzi, Prince Nyarko, Claudio Fronterre, Robert A. Cheke, Maria-Gloria Basáñez, Frances M. Hawkes

## Abstract

Progress towards elimination of onchocerciasis transmission is evaluated using entomological and serological indicators. The latter assesses anti-Ov16 IgG4 seroprevalence in children aged <10 years. A seroprevalence of <0.1% suggests that ivermectin Mass Drug Administration (MDA) can be stopped and post-treatment surveillance initiated, according to World Health Organization (WHO) guidelines. Surveillance of populations living in close proximity to vector breeding sites and first-line villages may mask spatial transmission heterogeneity. We mapped anti-Ov16 seroprevalence within a 25-km radius around a known *Simulium damnosum* s.l. breeding site in Asubende, Ghana, to assess its spatial patterns and their implications for serosurveillance.

A cross-sectional survey was conducted in September-November 2024 in 30 settlements. Individuals aged ≥3 years were recruited through convenience sampling. The Ov16 rapid diagnostic test (RDT) using dry blood spots was used to estimate overall and site-level seroprevalence. Generalized additive models were used to assess seroprevalence trends versus distance from the breeding site.

Among 2,479 participants with valid RDT results, overall seroprevalence was 10.0% (95% CI: 8.9%, 11.3%) and increased with age. Seroprevalence varied across sites (0–24.4%) and declined with increasing distance from the breeding site. Among 584 children <10 years old, seroprevalence was 1.5% (95% CI: 0.7%, 2.9%). Adjusting for RDT sensitivity and specificity, seroprevalence in this age-group was 0.7%, (95% CI: 0%, 2.4%). Seropositive children were detected up to 18 km from the breeding site, but none were identified at it.

The distance-related decrease in overall seroprevalence is consistent with spatial patterns in vector abundance at Asubende and implies higher cumulative exposure near the breeding site. The small number of children tested limited inference in this WHO target age-group. Detection of seropositive children away from, but not at, the breeding site highlights limitations of surveillance focused on first-line villages and supports broader spatial sampling to strengthen evidence for stop-MDA decisions.

**Author summary:** After decades of onchocerciasis control using ivermectin, many countries hope to demonstrate that they have interrupted transmission of this vector-borne parasitic disease using serological surveys. It is unclear whether exposure to infection is spatially clustered around the riverine breeding sites of the blackfly vectors and therefore whether this is where serosurveillance should focus. To address this, we collected dried blood spots from 2,480 consenting participants aged 3-96 years old from 29 communities within a 25-km radius of a known blackfly breeding site in Asubende, Ghana. A rapid diagnostic test was used to test the blood spots for antibodies to the *Onchocerca volvulus* parasite. We found that overall seropositivity declined with increasing distance from the breeding site, which suggests that cumulative exposure is higher near the breeding site, where vector population is also high. However, seropositivity in children (3–10-year-olds, n= 584), which is indicative of recent transmission, was spatially distributed across the study area and found up to 18 km from the breeding site. These findings are relevant to serosurveillance sampling strategies intended to detect low levels of transmission, which could miss peripheral evidence of ongoing transmission if they are focussed at breeding sites and those villages closest to them.

## Introduction

Onchocerciasis (river blindness) is a parasitic infection caused by the filarial nematode *Onchocerca volvulus* and transmitted among humans via the bites of female blackflies of the *Simulium* genus. Simuliid flies breed in fast-flowing, well-oxygenated water bodies such as rivers and rapids (1). Onchocerciasis is currently endemic in 28 countries in sub-Saharan Africa, two countries in South America and Yemen (2), and can lead to severe morbidity including dermatological conditions (3), blindness (4) and neuro-hormonal manifestations (5). Heavy infection load is associated with a significantly higher relative risk of mortality, particularly in the young (6). Decades of interventions, including vector control and ivermectin mass drug administration (MDA) targeting the parasite’s embryonic stages (the microfilariae, mainly responsible for the clinical manifestations) have substantially reduced the disease burden in affected regions. However, globally, approximately 20 million people were estimated to be infected in 2021 (7) and more than 250 million people still require treatment to interrupt onchocerciasis transmission (8). Interruption (elimination) of transmission is the goal proposed by the 2021–2030 World Health Organization (WHO) Roadmap on Neglected Tropical Diseases, which aims at 12 (approximately a third of) endemic countries being verified for interruption of transmission by 2030 (9). As of 2025, in the African region, only Niger has been verified for elimination of transmission (10), and Senegal is in the post-treatment surveillance phase (2). The main strategy advocated by the WHO is primarily annual MDA treatment with ivermectin. Semi-annual MDA may be used in high transmission areas or to accelerate interruption of transmission (9).

To evaluate progress toward elimination, specific entomological and epidemiological targets have been proposed. For entomological surveillance, the target is to achieve less than 0.05% of infectivity prevalence in blackfly vector samples of at least 6,000 flies collected throughout a transmission zone and tested for *O. volvulus* DNA in their heads. For serosurveillance, the WHO recommends testing for IgG4 antibodies to the *O. volvulus* Ov16 antigen in samples of 2000 children under 10 years old to determine if seroprevalence is at or below 0.1% (at the upper confidence interval bound) (11). According to current WHO guidelines, reaching this <0.1% threshold serves as an indicator of transmission interruption, supporting a recommendation to stop MDA (11). However, the WHO also recognises that the evidence provided by this measure has low certainty in its indication that transmission has been interrupted. This uncertainty is in part attributable to the diagnostic performance limitations of the available diagnostic tools for serology analysis, including enzyme-linked immunosorbent assays (ELISA) and Rapid Diagnostic tests (RDTs), which have a lower specificity than that required to reliably detect when seroprevalence is at or below 0.1% (12) (specificity should be at least 99.9%). Additionally, obtaining the appropriate sample size of children under 10 years old is operationally challenging (11, 12).

Despite these shortcomings, serological data provide a measure of cumulative exposure to *O. volvulus* infection, and may indicate ongoing transmission (13). This is particularly useful in low-prevalence or near-elimination settings, where entomological surveillance may be less reliable (e.g., due to very low vector population densities in the case of low prevalence, hypoendemic areas (14) or in areas where transmission conditions have been altered due to environmental change leading to declining vector populations (15)), which complicates estimation of transmission parameters. In these settings, serosurveillance may help in detecting ongoing transmission or potential infection resurgence, particularly if seropositivity is detected in younger individuals in the population.

Current conventions on onchocerciasis surveillance focus on sampling in first-line villages located at or near riverside vector breeding sites. However, previous research has demonstrated that blackflies can disperse several kilometres (km), including up to 500 km, from their breeding site (16-21). Furthermore, recent work by Kyomuhangi *et al.* in Benin and Ghana, documented spatial variation in blackfly biting rates within a 25-km radius around well-characterised breeding sites (Bétérou in Benin and Asubende in Ghana), as well as spatial and temporal heterogeneity in blackfly infectivity in the Benin study area (22). Together, these findings highlight the need for more spatially-extensive sampling strategies beyond breeding sites to better inform stop-MDA decisions.

As countries progress towards elimination and post-elimination phases and increasingly integrate serosurveillance with entomological surveillance to guide stop-MDA decisions, it is important to understand whether and how spatial heterogeneity in seroprevalence might inform sampling strategies for serological surveys.

Therefore, building on the entomological work reported in Kyomuhangi *et al.* (22), this study aims to map the spatial distribution of anti-Ov16 seroprevalence and explore how seroprevalence changes with distance from a known blackfly breeding site in Asubende in Ghana.

## Methods

This study was a cross-sectional serological survey conducted as part of a broader research project (22). The broader project investigated the spatial dispersal patterns of blackflies (*Simulium damnosum* sensu lato (s.l.)) in Benin and Ghana by assessing entomological indices at a primary breeding site and 40 surrounding data collection sites within a 25-km radius of the breeding site. The current study aimed to investigate spatial patterns of anti-Ov16 seroprevalence within the same area for Ghana. Consequently, the locations used in this study were determined based on the sampling framework established in the larger project (22).

### Study area

The study was conducted in and around Asubende, located in the Pru East district (Bono East region—part of the previous Brong-Ahafo region) of Ghana, between September and November 2024, during the region’s wet season. A breeding site along the Pru River (08°01’01.4"N, 00°58’53.8"W) was selected because of its history as a vector monitoring site under the WHO Onchocerciasis Control Programme in West Africa (OCP), with accessible historical data (capture point 1202). The main vector species in the area belong to the savannah members of the *Simulium damnosum* s.l. species complex (i.e., *S. damnosum* sensu stricto/*S. sirbanum*) (23). In the area, vector control started (by weekly aerial insecticidal larviciding of vector breeding sites) in the late 1980s as part of the OCP’s southern extension (24). In particular, around the Asubende focus, vector control activities began in 1986, but were suspended between 1987 and 1989 because of the first community trial of ivermectin being conducted in the area (25, 26). The focus was holoendemic at baseline, with a microfilarial prevalence of 80% and 72 microfilariae/skin snip (26). Larviciding was reinstated in 1990 and, according to the OCP, continued until 2001. Annual ivermectin MDA was implemented from 1987 (the first five years as part of the trial (26) until 2009), followed by biannual treatment from 2010 onwards (27). The therapeutic coverage during the trial ranged from 62% to 74% of the total population (26), increasing to 85–90% in 2011–2012 (27). By September-November 2024, when our study was conducted, the Asubende focus would have received 48 rounds of ivermectin MDA, comprised of 22 years of annual treatment (22 rounds) and 13 years of biannual treatment (26 rounds). Treatment was interrupted during 2020 because of the COVID-19 pandemic.

Within the larger project, a spatially-explicit radial sampling design was used to select 40 settlements within a 25-km radius of the breeding site (22). These settlements were selected on the basis that they had more than 100 inhabitants, as determined by using the Geo-Referenced Infrastructure and Demographic Data for Development (GRID3-SE) settlement extent dataset(22, 28), and the WorldPop project (29, 30). The selection process involved randomly selecting 30 settlements (within the 25-km radius), ensuring that they were at least 5 km apart, then randomly choosing 10 of these and including their nearest neighbouring settlements, resulting in a total of 40 collection sites.

In this study, only 29 of the original 40 collection sites were included, with 11 sites excluded (10 sites because field verification indicated they lacked resident human populations, and 1 site because it was inaccessible to the study team, as the road leading to it could only be accessed by motorcycle or bicycle). Figure 1 shows the breeding site and collection site locations selected for the study. In total, there were 30 collection sites, including the breeding site.

**Figure 1.**
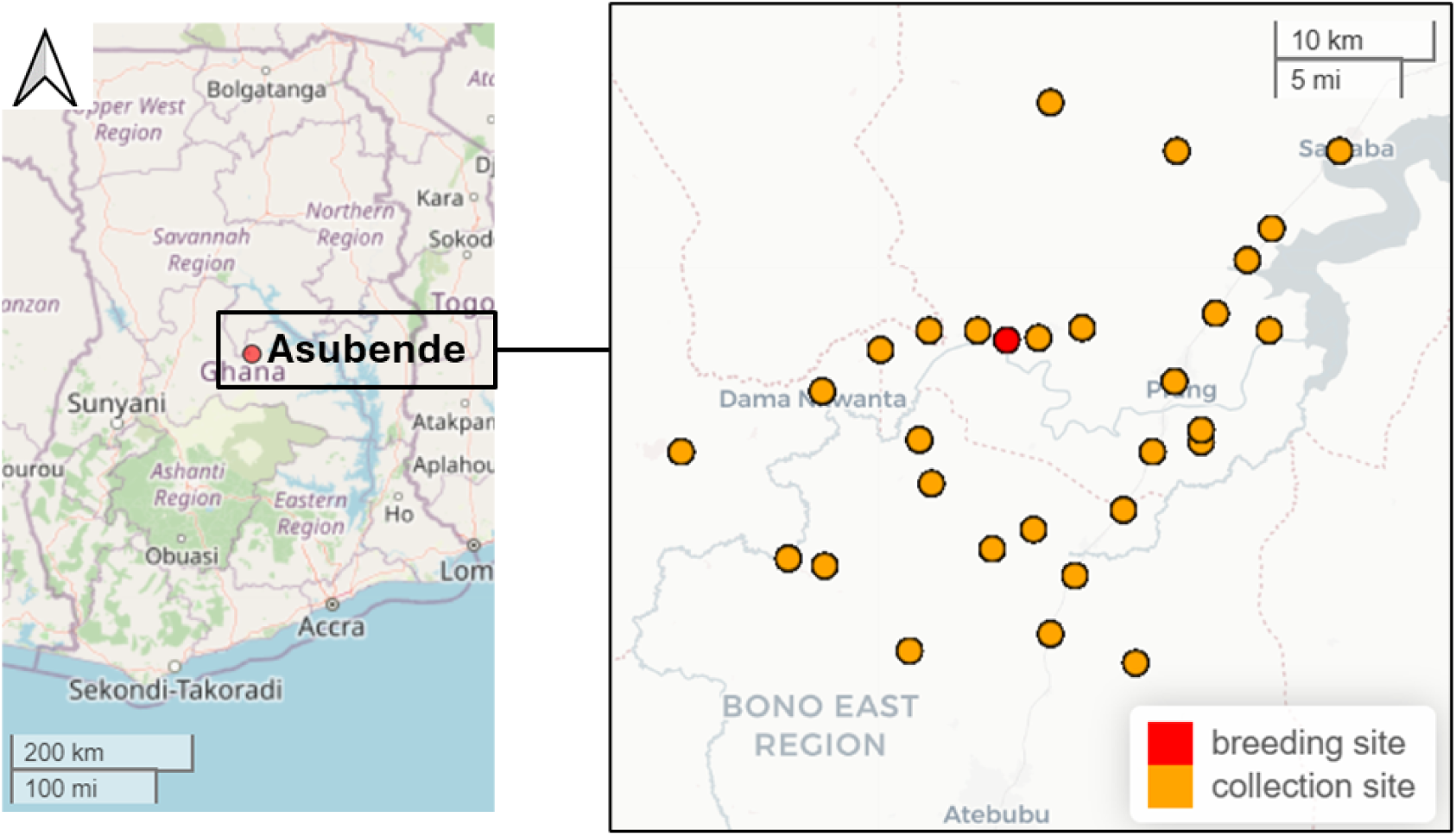
Study sites in Asubende, Ghana, in 2024. In the inset, the red circle represents the breeding site; the orange circles represent the remaining 29 collection sites, which were within a 25-km radius of the breeding site. The grey lines and shapes indicate water bodies.

### Recruitment of participants

Recruitment of participants to the study was done through convenience sampling. The recruitment period started on the 1^st^ of September 2024 and ended on the 14^th^ of November 2024. To prepare for the survey, announcements were made in the study site communities approximately one to two weeks in advance of data collection. These announcements were delivered through the community information centres and by using the traditional gong-gong method, where a local gong was periodically sounded through the community to attract attention, as previously described (31). Residents were invited to gather at designated community centres where the purpose of the study was explained first in English, then translated into the local Twi language, in the presence of village elders and community members. All volunteers aged 3 years and older were invited to participate in the study. Participants were recruited once written informed consent was obtained; for children aged 3-17 years, consent was provided by a parent or guardian, with an additional signed assent form provided by children aged 10-17 years.

### Serology data

Blood samples were collected from each participant via finger-prick using a sterile lancet or needle. The blood from the puncture was then transferred onto a Whatman 903™ protein saver card to prepare five dried blood spots (DBS) per participant. The spots were prepared by uniformly saturating the entire circle of the card through quick and gentle touches (not pressing) of the puncture site to the filter paper. The DBS samples were labelled with the participant’s unique identifier and the collection date, and allowed to air dry in a clean, dry area away from direct sunlight for 30 minutes. Once fully dry, each card was placed into a gas-impermeable plastic bag containing a desiccant pack. The bags were sealed and stored in a dry box for transportation to the laboratory, where they were kept at -20°C until analysis.

Ov16 RDTs (SD BIOLINE Onchocerciasis IgG4 RDT, Abbot, Republic of Korea) were used to test for antibody seropositivity to the Ov16 *O. volvulus* antigen in the DBS eluates using a previously described protocol (32). RDT results read after 24 hours were used for analysis. The Ov16 RDT with DBS has been determined to have 80% sensitivity and 99% specificity (33).

### Exposure data

During the serology data collection, participants aged 11 years and older were additionally invited to take part in interviews using a previously designed and tested questionnaire (34). This questionnaire gathered information on participants’ sociodemographic characteristics, as well as exposure variables (to infection, ivermectin, bites) such as: duration of residency within the community, ivermectin intake, interaction with rivers or streams, and their experiences of the location, timing and intensity of blackfly bites. The interviews were administered in English and Twi by trained members of the study team, and participants’ responses were recorded on a custom-built application using KoboToolbox (35).

### Analysis

All analyses were conducted in the R programming language (version 4.5.1) (36). For the descriptive analyses, seroprevalence estimates and their 95% confidence intervals (CIs) were calculated using Clopper-Pearson, exact 95% CIs. Exploratory analyses investigating trends between outcome and explanatory variables were conducted using Generalized Additive Models (GAMs). Graphs were generated using the ggplot2 (37) package in R, and maps were created using the leaflet R package (38).

## Results

A total of 2,480 individuals participated, with sample sizes ranging from 26 to 117 individuals per site. Overall, 59.2% of participants were female while 40.8% were male. Participants’ ages ranged from 3 to 96 years, with a median age of 25 years. Figure 2a shows the age distribution of the sampled population.

**Figure 2.**
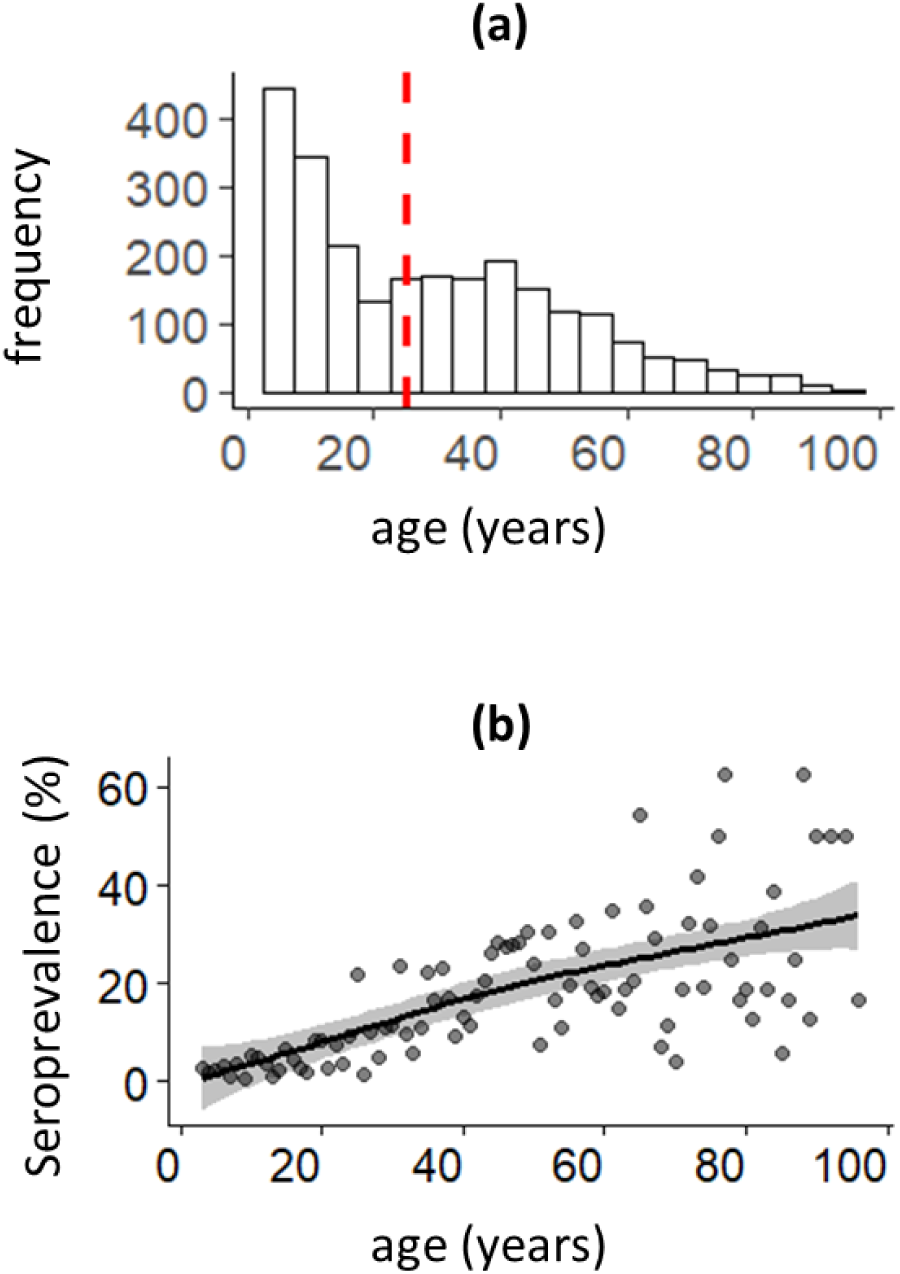
Age distribution of the sampled population and anti-Ov16 seroprevalence vs. age at the 30 sites. (a) Age distribution of the 2,480 study participants. The red line indicates the median age (25 years). (b) Anti-Ov16 seroprevalence across age for the 2,479 participants with valid Ov16 rapid diagnostic test (RDT) (using dried blood spots (DBS)) results. The dark grey circles show the observed seroprevalence estimates; the black smoothed curve represents the age-specific trend estimated using a binomial generalized additive model (GAM); the shaded grey area indicates the 95% CI around the model.

Out of the 2,480 RDTs performed, one test was void. Among all participants with valid RDT results (2,479), 249 were seropositive, with an overall anti-Ov16 seroprevalence equal to 10.0% (95% CI: 8.9%, 11.3%). Seroprevalence increased with age (Figure 2b). Among 584 children under 10 years of age across all sites, 9 were seropositive (2 boys aged 3 years; 3 girls aged 4-5; 2 boys aged 6, and 1 girl and 1 boy aged 8), with an anti-Ov6 seroprevalence of 1.5% (95% CI: 0.7%, 2.9%). Considering the 80% sensitivity and 99% specificity of the Ov16 RDT with DBS (33), the ‘adjusted’ seroprevalence and adjusted 95% CIs (39) in children aged <10 years would be equal to (seroprevalence + 0.99 – 1) / (0.80 + 0.99 –1) = 0.0068 (0.7%, 95% CI: 0%, 2.4%).

Seroprevalence varied spatially across sites, ranging from 0 to 24.4% (Figure 3a; Table in S1 Table). However, overall, seroprevalence tended to decline with increasing distance from the breeding site (Figure 3b).

**Figure 3.**
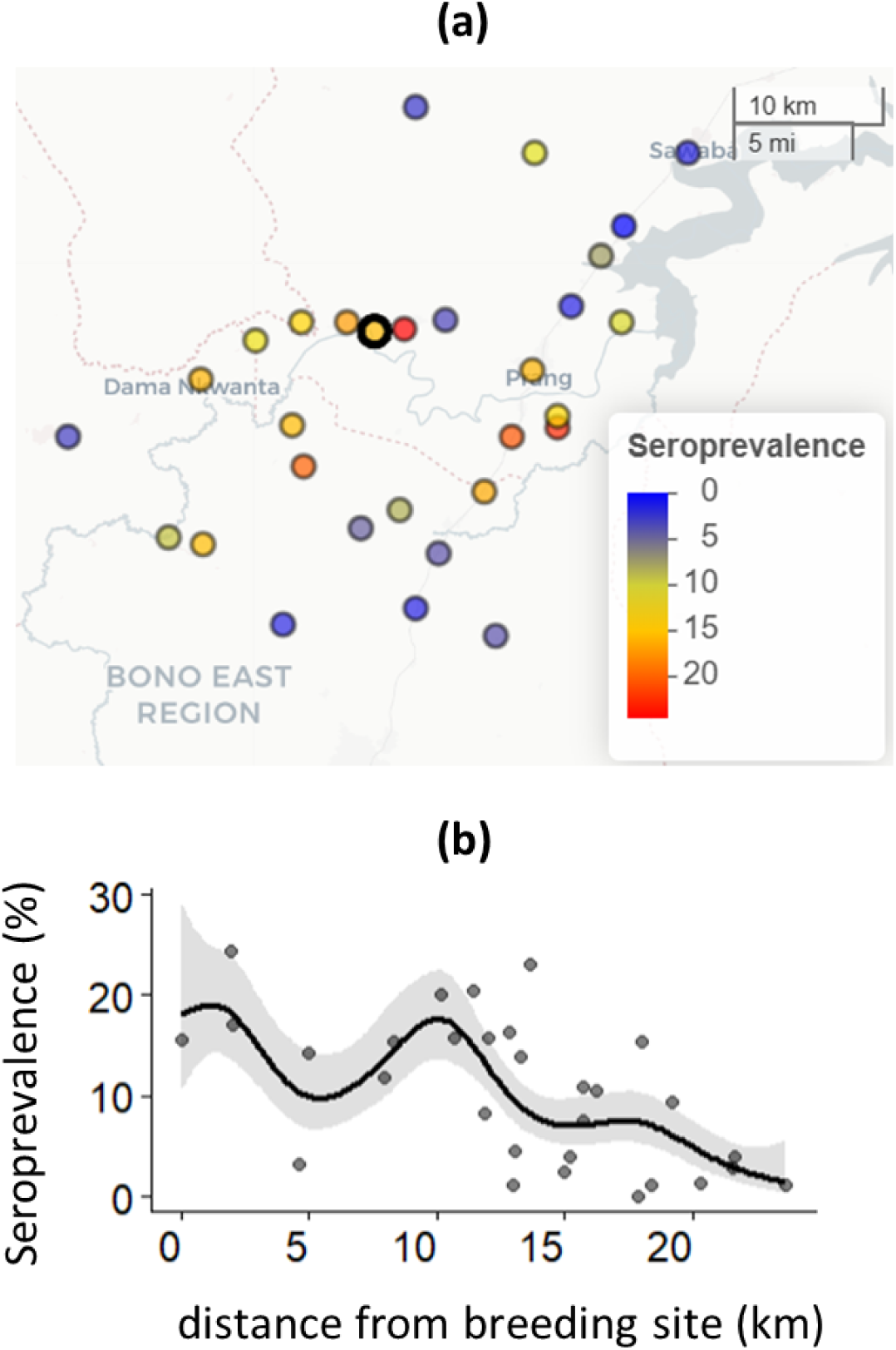
Spatial distribution of estimated anti-Ov16 seroprevalence and trend in seroprevalence vs distance from breeding site. (a) Map showing the estimated seroprevalence among the 2,479 participants with valid Ov16 RDT (using DBS) results across the 30 study sites, with the breeding site highlighted by a thick black border. (b) Trend in overall anti-Ov16 seroprevalence by distance from the breeding site. The dark grey circles represent the observed seroprevalence estimates (in those age 3–96 years) at each samples site; the black smoothed curve shows the trend estimated using a binomial generalized additive model (GAM); the grey shaded area is the 95% CI around the model.

The number of children under 10 years of age per site was small, ranging from 1 to 38 (Table in S1 Table). Only 7 out of the 30 sites (23.3%) recorded at least one seropositive child in this age group (Figure 4), with site-level seroprevalence ranging from 0 to 13.0%, the latter at approximately 16 km from the breeding site (Table in S1 Table). Notably, no seropositive children were identified at the breeding site, whereas a seropositive child was detected up to 18 km away.

**Figure 4.**
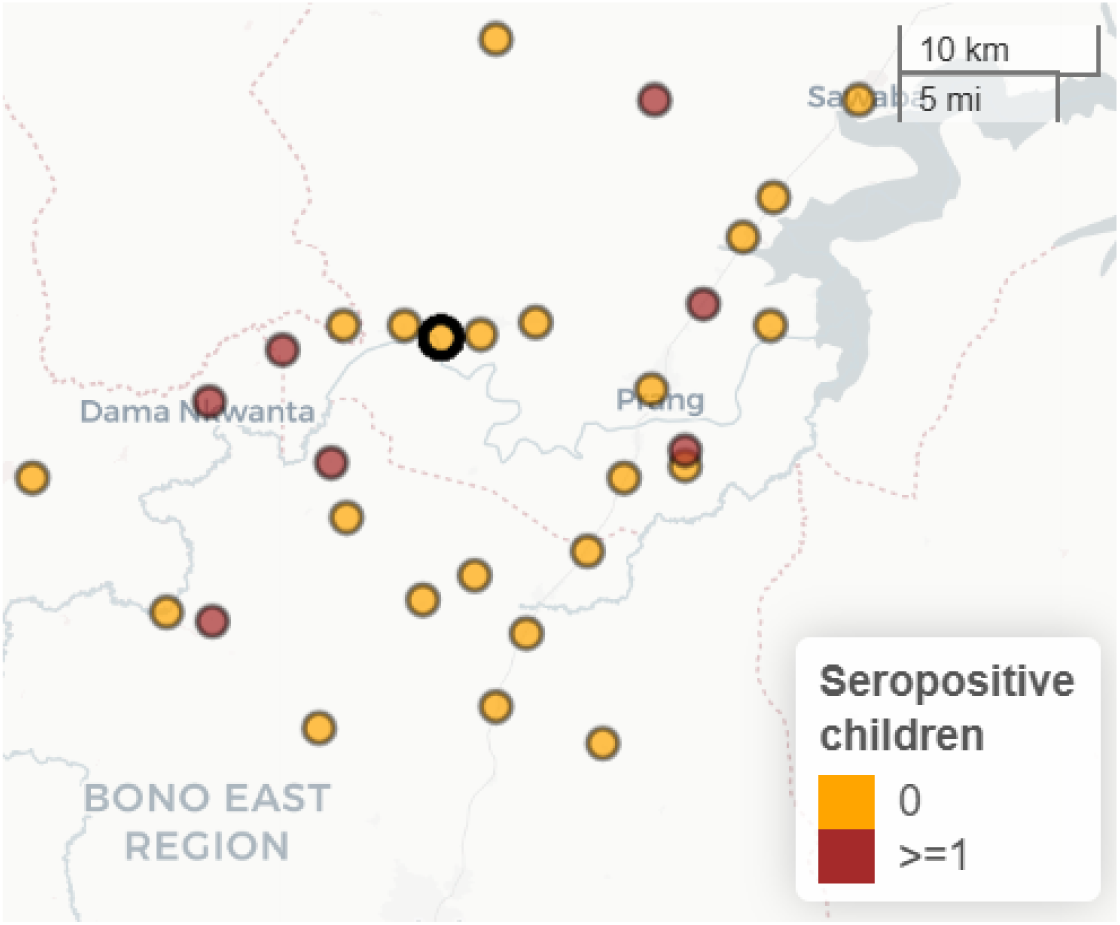
Map showing which sites had at least one anti-Ov16 seropositive child under 10 years old. The breeding site is marked with a thick black border. Red circles indicate where there were one or more seropositive children, while yellow circles show where there were no seropositive children.

Out of the 2,479 participants with a valid RDT result, questionnaire-derived exposure data were available for 1,358 individuals (54.8%). Among participants without exposure data, 57.3% were under 11 years old and thus ineligible for the interviews. The remaining 42.7% lacked exposure data mainly because participants had difficulty understanding the questions (particularly non-adult participants), and in certain instances, language barriers hindered the completion of interviews (specifically, in 7 out of 30 sites, exposure data were not collected due to language issues).

Among the 1,358 participants with available exposure data, most reported similar levels of exposure across various indicators. The majority had lived in the village for an extended period, with 78.9% residing in their villages for over 11 years. Additionally, the vast majority (98.9%) had a stream or river nearby, and 56.5% visited the stream or river daily (Table in S2 Table).

Ivermectin intake was consistently high, with 97.1% having previously taken ivermectin, 83.5% having taken ivermectin five or more times overall, and 77.2% having taken ivermectin in the last round of MDA, which took place in July 2024 (Table in S2 Table).

Regarding the location and timing of blackfly bites, only 5.8% of participants reported that they most often experience bites at home, while the majority (87.8%) indicated that most bites occur at the riverside or on a farm. Furthermore, most participants (73.0%) stated that blackfly biting is most frequent during the wet season, primarily from August to October (Figure in S3 Figure), which coincides with the data collection for this study. Regarding their experiences over time, 67.5% of respondents observed a decline in blackfly biting in recent years (Table in S2 Table).

Because reported exposure variables showed little variation across participants, no significant associations with seropositivity were detected (Table in S2 Table). Consequently, these variables were unlikely to meaningfully modify the relationship between seroprevalence and distance from the breeding site and were not used for further analysis.

## Discussion

This study mapped the spatial distribution of anti-Ov16 seroprevalence across different sites located within a 25-km radius of a known blackfly breeding site in Asubende, Ghana, and examined the relationship between seroprevalence and distance from the breeding site. Results showed a clear spatial gradient in seroprevalence across the sampled area as overall seroprevalence in the general population tended to decline with increasing distance from the breeding site.

This pattern is consistent with entomological findings from the broader project described by Kyomuhangi *et al.* which showed that blackfly abundance declined with increasing distance from breeding sites (22). It therefore supports the expectation that cumulative exposure to infection is highest near breeding sites and decreases with distance. The observed spatial heterogeneity in seroprevalence across sites further mirrors the entomological patterns.

Although overall anti-Ov16 seroprevalence tended to decline with distance from the breeding site, this pattern was not observed among children under 10 years of age—the group prioritised by the WHO for serological surveillance. This discrepancy is probably due to the low numbers of children sampled and the very low number of seropositive children, as the seroprevalence in this age group was 1.5%, and 23 of 30 sites recorded zero cases. The high proportion of zero-seroprevalence sites limits the ability to detect spatial trends in seroprevalence for this age group. These findings are consistent with the very low blackfly catches reported for 2023–2024 (22) and participant interviews in this current study describing reduced blackfly biting in recent years. However, at the Asubende breeding site, daily biting rates of 119/person (2023) and 47/person (2024) were recorded by human landing catches, which when multiplied by 365 to obtain a crude approximation to the annual biting rate, would yield values of 44,000 (2023) and of 17,000 (2024) bites/person/year (22). These values are well within the range of 5,000–45,000 recorded by the OCP at the 1202 (Asubende) capture point prior to vector control (between 1978 and 1985), and also in line with the 20,000–31,000 bites/person/year recorded between 2010 and 2016 (8–14 years after cessation of vector control) in villages of the Asubende focus (Francis V.D. Veriegh, pers. comm.), suggesting that vector biting rates may have bounced back to their original levels (23).

Although we sampled at 30 sites, only 584 children were recruited to the study, well below the 2,000 per focus or transmission zone recommended by the WHO for serosurveillance (11). This highlights the logistical challenges of implementing this serosurveillance approach for meaningful policy decisions. Notably, no seropositive children under 10 years of age were identified at the breeding site itself, while those that were seropositive were scattered throughout the study area, suggesting that surveillance restricted to breeding sites could miss peripheral evidence of ongoing transmission. Such an approach may therefore underestimate the epidemiological situation in surrounding communities and risk premature stop-MDA decisions. The exposure data, collected from participants aged 11 and above, showed remarkably similar reported exposure indicators across sites: long-term residency, frequent river contact, and high reported ivermectin intake. This homogeneity probably reflects the relatively small spatial scale of the study and suggests broadly similar exposure opportunities for this age group across the 30 sites. However, we did not collect comparable exposure information for children under 10, so we cannot rule out heterogeneity in exposure within this younger age group or differences between children and older participants. Consequently, some of the variation (or lack thereof) in under-10 seroprevalence might be attributable to unmeasured differences in exposure among children. Furthermore, there were challenges in completing the questionnaire by all those invited to take part due to language barriers (although translators were used), and the study was based on convenience (rather than a probability-based) sampling. Therefore, it is not possible to discount the possibility that individuals volunteering to participate in the study would also be those who agree to take ivermectin regularly, in addition to potential social desirability bias (34).

The performance of the currently recommended antibody diagnostic tools (33) constitutes a limitation of any anti-Ov16 seroprevalence study. Specifically, current RDTs do not have the appropriate sensitivity and specificity required for serosurveillance as recommended by the WHO (12). When seroprevalence in the target age group is close to zero, specificity and sensitivity of diagnostic tools can materially affect inferences about exposure and transmission. In near-elimination settings—and particularly where under-10 seropositivity is rare—highly sensitive, and more importantly, highly specific diagnostics are critical, the former to avoid false negatives that could mask exposure/transmission, and the latter to minimise false positives that would trigger unnecessary programmatic responses (40). When data from all sites were combined, anti-Ov16 seroprevalence in the under 10-year-olds was 1.5% (95% CI = 0.7%, 2.9%), and after accounting for the sensitivity and specificity of the Ov16 RDT with DBS (33), the ‘adjusted’ seroprevalence was 0.7%, (95% CI: 0%, 2.4%). Given that WHO guidelines permit only one seropositive child at the prescribed sample size, the identification of nine cases in a substantially smaller sample suggests that, had the recommended sampling strategy been feasible and a larger sample size been examined, the upper confidence limit would probably have exceeded the 0.1% anti-Ov16 seroprevalence threshold for ongoing transmission. Given that the average period for detectable IgG4 antibody production is 15 (10-18 months), coinciding with that for detectable skin microfilariae, the seropositive three-year-olds in our study were probably exposed to sufficient transmission between 2020 (their earliest possible birth year) and mid-2024 to have acquired worms of both sexes capable of initiating an established infection eliciting anti-IgG4 seropositivity (13, 41). For programmatic decision-making, these findings would need to be confirmed using the recommended sampling strategy and sample sizes.

## Conclusion

Anti-Ov16 seroprevalence tended to decline with increasing distance from the breeding site, consistent with entomological evidence of declining vector abundance with distance and highlighting spatial heterogeneity in exposure and transmission. However, patterns in children aged under 10 years were not conclusive, owing to very low seroprevalence in this age group, combined with low sample size. The absence of seropositive children at the breeding site, alongside detection of seropositive cases away from the breeding site, suggests that surveillance limited to first-line villages may misrepresent the epidemiological situation more broadly around the focus. Results emphasize the need for broader serological sampling around breeding sites, intensified R&D investment in highly sensitive and specific diagnostics appropriate to reliably measure proposed seroprevalence thresholds for supporting stop-MDA decisions, and continued work to scrutinise such thresholds as indicative of interruption of transmission (42).

## Data and software availability

All raw data (excluding precise GPS coordinates) are available in the Supporting Information. Exact spatial data are withheld due to ethical and privacy considerations. Data sufficient to reproduce all analyses, including derived distance measures, are provided in the public dataset. Access to precise GPS coordinates may be granted upon reasonable request to the corresponding authors, subject to review of the proposed research purpose and completion of an appropriate data-sharing agreement. Where applicable, applicants may be required to provide evidence of institutional ethical approval. Data will be shared in accordance with relevant ethical and data protection requirements.

## Authors’ contributions

Conceptualisation: IK, KBO, CF, RAC, M-GB, FMH. Methodology: IK, KBO, AA, DKO, P-CK, PN, CF, RAC, M-GB, FMH. Investigation: KBO, AA, DKO, P-CK, PN, Validation: KBO, AA, DKO, P-CK, PN.

Software: IK, CF. Formal analysis: IK, CF, KBO, MG-B. Data curation: IK. Writing – original draft: IK. Writing – review and editing: KBO, AA, DKO, P-CK, PN, CF, RAC, M-GB, FMH. Visualization: IK. Supervision: KBO, FMH. Project administration: FMH. Funding acquisition: IK, KBO, CF, RAC, M-GB, FMH.

## Acknowledgements

We are grateful to the study participants and the communities in Ghana for their time, cooperation, and willingness to take part in this research. We also extend our appreciation to the study team at the Consortium for Neglected Tropical Diseases and One Health at the University of Energy and Natural Resources in Ghana for their dedication and invaluable contributions to the project.

## Ethics

Ethics approvals were obtained from the Committee for Human Research and Ethics of the University of Energy and Natural Resources in Ghana (ref: CHRE/AP/241/024) and the University of Greenwich Research Ethics Board (ref: UREB 23.4.5.2).

## Supporting information

**S1 Table. Seroprevalence estimates from the different sites among all participants with a valid RDT result, and seroprevalence among participants under 10 years of age.** Data are ordered according to distance from the breeding site.

**S2 Table. Participants’ responses to exposure (infection, ivermectin) questionnaire.** The data were collected during individual interviews with 1,358 participants. Binomial GAMs were conducted to explore the relationship between seropositivity and several exposure variables. The models specify seropositivity as the binary outcome variable, and the respective categorical exposure variable as the explanatory variable.

**S3 Figure. The percentage of participants who selected the month response to the question: ‘Is there a certain month that the biting by blackflies is typically worst?’**

## References

1. Burnham G. Onchocerciasis. Lancet. 1998;351(9112):1341–6.

2. World Health Organization. Status of endemicity of onchocerciasis. Geneva: World Health Organization (last updated 07 October 2025). Available from: https://www.who.int/data/gho/data/indicators/indicator-details/GHO/status-of-endemicity-of-onchocerciasis

3. Murdoch M, Hay R, Mackenzie C, Williams J, Ghalib H, Cousens S, et al. A clinical classification and grading system of the cutaneous changes in onchocerciasis. Br J Dermatol. 1993;129(3):260–9.

4. Hopkins A, Boatin BA. Onchocerciasis. In: Selendy JMH, editor. Water and sanitation-related diseases and the environment: challenges, interventions, and preventive measures. 1^st^ ed. Hoboken (NJ): John Wiley & Sons; 2011. p. 133-49. Available from: 10.1002/9781118148594.ch11.

5. Van Cutsem G, Siewe Fodjo JN, Hadermann A, Amaral LJ, Trevisan C, Pion S, et al. Onchocerciasis-associated epilepsy: Charting a path forward. Seizure. 2026;135:105–14.

6. Walker M, Little MP, Wagner KS, Soumbey-Alley EW, Boatin BA, Basanez M-G. Density-dependent mortality of the human host in onchocerciasis: relationships between microfilarial load and excess mortality. PLOS Negl Trop Dis. 2012;6(3):e1578.

7. GBD 2021 Diseases and Injuries Collaborators. Global incidence, prevalence, years lived with disability (YLDs), disability-adjusted life-years (DALYs), and healthy life expectancy (HALE) for 371 diseases and injuries in 204 countries and territories and 811 subnational locations, 1990–2021: a systematic analysis for the Global Burden of Disease Study 2021. Lancet. 2024;403(10440):2133-61.

8. World Health Organization. Elimination of human onchocerciasis: progress report, 2023–2024. Geneva: World Health Organization; 2024. Available from: https://www.who.int/publications/i/item/who-wer-9941-577-590.

9. World Health Organization. Ending the neglect to attain the sustainable development goals: a rationale for continued investment in tackling neglected tropical diseases 2021–2030. Geneva: World Health Organization; 2022. Available from: https://www.who.int/publications/i/item/9789240010352.

10. World Health Organization. WHO verifies Niger as the first country in the African Region to eliminate onchocerciasis [Internet]. Geneva: World Health Organization; 2025 Jan 30. Available from: https://www.who.int/news/item/30-01-2025-who-verifies-niger-as-the-first-country-in-the-african-region-to-eliminate-onchocerciasis.

11. World Health Organization. Guidelines for stopping mass drug administration and verifying elimination of human onchocerciasis: criteria and procedures. Geneva: World Health Organization; 2016 (updated 2020). Available from: https://www.who.int/publications/i/item/9789241510011. Report No.: 9241510013.

12. Gass KM. Rethinking the serological threshold for onchocerciasis elimination. PLOS Negl Trop Dis. 2018;12(3):e0006249.

13. Basáñez M-G, Ramani A, Stapley J, Dixon M, Hamley J, Amaral L, et al. Modelling anti-Ov16 Seroprevalence for the Control and Elimination of Onchocerciasis [preprint]. Available from: 10.21203/rs.3.rs-7140160/v1. 2025.

14. Stapley JN, Hamley JI, Basáñez M-G, Walker M. Modelling transmission thresholds and hypoendemic stability for onchocerciasis elimination. PLOS Comput Biol. 2025;21(4):e1013026.

15. Post RJ, Laudisoit A, Mandro M, Lakwo T, Laemmer C, Pfarr K, et al. Identification of the onchocerciasis vector in the Kakoi-Koda focus of the Democratic Republic of Congo. PLOS Negl Trop Dis. 2022;16(11):e0010684.

16. Garms R, Walsh J, Davies J. Studies on the reinvasion of the Onchocerciasis Control Programme in the Volta River Basin by *Simulium damnosum* s.I. with emphasis on the south-western areas. Tropenmed Parasitol. 1979;30(3):345–62.

17. Gibbins E. Uganda Simuliidae. Trans R Entomol Soc Lond. 1936;85:217–42.

18. Le Berre R. Contribution à l’étude biologique et écologique de *Simulium damnosum* Theobald, 1903 (Diptera, Simuliidae). Mém ORSTOM. 1966;17:1–204.

19. Germain M, Eouzan JP, Ferrara L. Données sur les facultés de dispersion de deux diptères d’intérêt médical: *Aedes africanus* (Theobald) et *Simulium damnosum* Theobald, dans le domaine montagnard du nord du Cameroun occidental. Cah ORSTOM Sér Entomol Med Parasitol. 1972;10:291–300.

20. Duke BO. The differential dispersal of nulliparous and parous *Simulium damnosum*. Tropenmed Parasitol. 1975;26(1):88–97.

21. Renz A, Wenk P. Studies on the dynamics of transmission of onchocerciasis in a Sudan-savanna area of North Cameroon I: Prevailing *Simulium* vectors, their biting rates and age-composition at different distances from their breeding sites. Ann Trop Med Parasitol. 1987;81(3):215–28.

22. Kyomuhangi I, Carnaghi M, Boko-Collins P, Soglongbe BB, Koukpo ZC, Otabil KB, et al. Spatial patterns of blackfly (*Simulium damnosum s.l*.) dispersal in Ghana and Benin and their implications for onchocerciasis surveillance. VeriXiv [preprint]. 2026;3:116. doi:10.12688/verixiv.3029.1.

23. Lamberton PHL, Cheke RA, Walker M, Winskill P, Osei-Atweneboana MY, Tirados I, et al. Onchocerciasis transmission in Ghana: biting and parous rates of host-seeking sibling species of the *Simulium damnosum* complex. Parasit Vectors. 2014;7(1):511.

24. O’Hanlon SJ, Slater HC, Cheke RA, Boatin BA, Coffeng LE, Pion SD, et al. Model-based geostatistical mapping of the prevalence of *Onchocerca volvulus* in West Africa. PLOS Negl Trop Dis. 2016;10(1):e0004328.

25. Remme J, Baker R, De Sole G, Dadzie K, Walsh J, Adams M, et al. A community trial of ivermectin in the onchocerciasis focus of Asubende, Ghana. I. Effect on the microfilarial reservoir and the transmission of Onchocerca volvulus. Trop Med Parasitol. 1989;40(3):367–74.

26. Alley E, Plaisier A, Boatin B, Dadzie K, Remme J, Zerbo G, et al. The impact of five years of annual ivermectin treatment on skin microfilarial loads in the onchocerciasis focus of Asubende, Ghana. Trans R Soc Trop Med Hyg. 1994;88(5):581–4.

27. Frempong KK, Walker M, Cheke RA, Tetevi EJ, Gyan ET, Owusu EO, et al. Does increasing treatment frequency address suboptimal responses to ivermectin for the control and elimination of river blindness? Clin Infect Dis. 2016;62(11):1338–47.

28. Center for International Earth Science Information Network (CIESIN), Foundation F, Fund UP, WorldPop UoS. Mapping and classifying settlement locations. Palisades (NY): Columbia University; 2021. Available from: https://eprints.soton.ac.uk/469540/1/GRID3_Settlement_White_Paper_June2021.pdf.

29. Tatem AJ, Noor AM, Von Hagen C, Di Gregorio A, Hay SI. High resolution population maps for low income nations: combining land cover and census in East Africa. PLOS One. 2007;2(12):e1298.

30. Linard C, Gilbert M, Snow RW, Noor AM, Tatem AJ. Population distribution, settlement patterns and accessibility across Africa in 2010. PLOS One. 2012;7(2):e31743.

31. Otabil KB, Basáñez MG, Ankrah B, Opoku SA, Kyei DO, Hagan R, et al. Persistence of onchocerciasis and associated dermatologic and ophthalmic pathologies after 27 years of ivermectin mass drug administration in the middle belt of Ghana. Trop Med Int Health. 2023;28(11):844–54.

32. Peck RB, Golden AL. Assay of dried blood spots using the SD BIOLINE Onchocerciasis IgG4 Rapid Test [Internet]. Seattle (WA): PATH; 2019. Available from: https://www.path.org/our-impact/resources/assay-dried-blood-spots-using-sd-bioline-onchocerciasis-igg4-rapid-test/.

33. World Health Organization. Onchocerciasis elimination mapping: a handbook for national elimination programmes. Geneva: World Health Organization; 2024. Available from: https://www.who.int/publications/i/item/9789240099227.

34. Otabil KB, Basáñez M-G, Ankrah B, Bart-Plange EJ, Babae TN, Kudzordzi P-C, et al. Non-adherence to ivermectin in onchocerciasis-endemic communities with persistent infection in the Bono Region of Ghana: a mixed-methods study. BMC Infect Dis. 2023;23(1):805.

35. Lakshminarasimhappa M. Web-based and smart mobile app for data collection: Kobo Toolbox/Kobo collect. J Indian Libr Assoc. 2022;57(2):72–9.

36. R Core Team. R: A language and environment for statistical computing [software]. Vienna: R Foundation for Statistical Computing; 2021. Available from: https://www.R-project.org/. 2021.

37. Wickham H. ggplot2: elegant graphics for data analysis. New York: Springer; 2016.

38. Cheng J, Schloerke B, Karambelkar B, Xie Y, Aden-Buie G. leaflet: create interactive web maps with the JavaScript ‘Leaflet’ library [software]. Vienna: R Foundation for Statistical Computing; 2025. Available from: https://rstudio.github.io/leaflet/.

39. Rogan WJ, Gladen B. Estimating prevalence from the results of a screening test. Am J Epidemiol. 1978;107(1):71–6.

40. Otabil KB, Basáñez M-G, Ameyaa E, Oppong M, Mensah P, Gyasi-Ampofo R, et al. Usability, acceptability, and cost of the SD BIOLINE Ov16 rapid diagnostic test for onchocerciasis surveillance in endemic communities in the middle belt of Ghana. PLOS Negl Trop Dis. 2025;19(8):e0012191.

41. Cama VA, McDonald C, Arcury-Quandt A, Eberhard M, Jenks MH, Smith J, et al. Evaluation of an OV-16 IgG4 Enzyme-Linked Immunosorbent Assay in Humans and Its Application to Determine the Dynamics of Antibody Responses in a Non-Human Primate Model of *Onchocerca volvulus* Infection. Am J Trop Med Hyg. 2018;99(4):1041–8.

42. Hamley JI, Walker M, Coffeng LE, Milton P, de Vlas SJ, Stolk WA, et al. Structural uncertainty in onchocerciasis transmission models influences the estimation of elimination thresholds and selection of age groups for seromonitoring. J Infect Dis. 2020;221(Supplement_5):S510-S8.

